# Vaccine efficacy for COVID-19 outbreak in New York City

**DOI:** 10.1101/2021.12.18.21268024

**Authors:** J. Demongeot, Q. Griette, P. Magal, G. F. Webb

**Affiliations:** Université Grenoble Alpes, AGEIS EA7407, F-38700 La Tronche, France; Univ. Bordeaux, IMB, UMR 5251, F-33400 Talence, France. CNRS, IMB, UMR 5251, F-33400 Talence, France; Mathematics Department, Vanderbilt University, Nashville, TN, United States

**Keywords:** Vaccine efficacy, Corona virus, reported and unreported cases, parameters identification, epidemic mathematical model

## Abstract

This article aims to study the COVID-19 data for New York City. We use both the daily number of second does vaccination and the daily number of reported cases for New York City. This article provides a method to combine an epidemic model and such data. We explore the influence of vaccine efficacy on our results.

## 1 Introduction

Developing vaccines against an infectious agent often requires years of research and testing to ensure efficacy and safety. In contrast, in the case of COVID-19, the vaccines took less than a year to develop and deploy. This rapid development has left many open questions whose answers may affect the usefulness of the epidemiological models proposed for COVID-19. In particular, vaccination efficacy rates are different for different populations in terms of the level and duration of vaccination immunization. We will study vaccinated efficacy according to the epidemic state of vaccinated individuals: susceptible, infected, or having received vaccine doses.

We will neglect both the duration of the immunization period (we will assume it is permanent) and the length of the acquisition phase of immunization after vaccine injection (we will assume it is identical for all vaccinated individuals). We will suppose the vaccination efficacy independent of age while noting that this hypothesis is very restrictive.

More careful considerations encourage caution with regard to the above simplifications and could lead to further studies. For example, vaccination efficacy is dependent on the appearance of variants and the existence of cross-immunization:

1. Inducing an antibody response may help select variants [25], a phenomenon very difficult to control because it is impossible to know what exact dose of antigenic Spike protein is released after each vaccination and what is its pharmacokinetics and its bio-distribution over time.
2. In the development of mRNA vaccines, cross-immunity was overlooked entirely [19]. There are anti-coronavirus antibodies and many epitopes common to the various endemic known coronaviruses, conserved with SARS-CoV-2. Vaccination ignores pre-existing cross-immunity, which is unfortunate, as the doses injected could be adjusted for a response via cross-immunity against epitopes common to coronaviruses. Young individuals are those whose cross-immunity is still active, and it would be helpful to design a vaccination policy to obtain the best efficacy per target population at risk.

Despite these limitations, a first simplifying approach can lead to a model making it possible to predict effective vaccination coverage at the population level that will prevent the appearance of successive epidemic waves.

Daily vaccination data, even if they are global and unrefined (for example, by age group or social classification), make it possible to better understand the effect of vaccination policy and test the consequences of changes in this policy to improve effectiveness. We use an epidemic model to understand the complex interactions between the epidemic dynamic and the epidemic data. Our model considers the changes in the public health policy, such as confinement, social distancing measures, etc., through the time-dependent transmission rate in the model. Data consist of the daily number of reported cases and the daily number of second doses of vaccine. We refer to [12, 13, 14, 15, 16, 18, 22, 23] for more results on the subject.

In the study, we propose a new model for vaccination implementation. We can connect the model with vaccination to a model without vaccination. We will find a simple transformation for the epidemic data to combine the daily reported case data and the cumulative number of vaccinated individuals.

We can use the model to explore controlling the dynamics of virus propagation, for example, by rapidly slowing down an epidemic wave. In this new model, we will take explicitly into account the variable corresponding to the size of the vaccinated population. We will simulate increasing efficacy of vaccination scenarios. We will apply our model to the COVID-19 epidemic in New York City.

## 2 Materials and Methods

### 2.1 Data

The data are taken from the New York City Department of Health and Mental Hygiene [26]. The epidemic of Sars-CoV2 started in NYC on February 29, 2020. The first complete vaccination (i.e., vaccination with two doses) started in NYC on December 15, 2020. In Figure 1-(a), the green dots correspond to the day by day constant values of the function CR_Data_(*t*)′ that is used in the model. In Figure 1-(b), this green curve corresponds to the value function 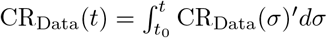 that is used in the model.

**Figure 1:**
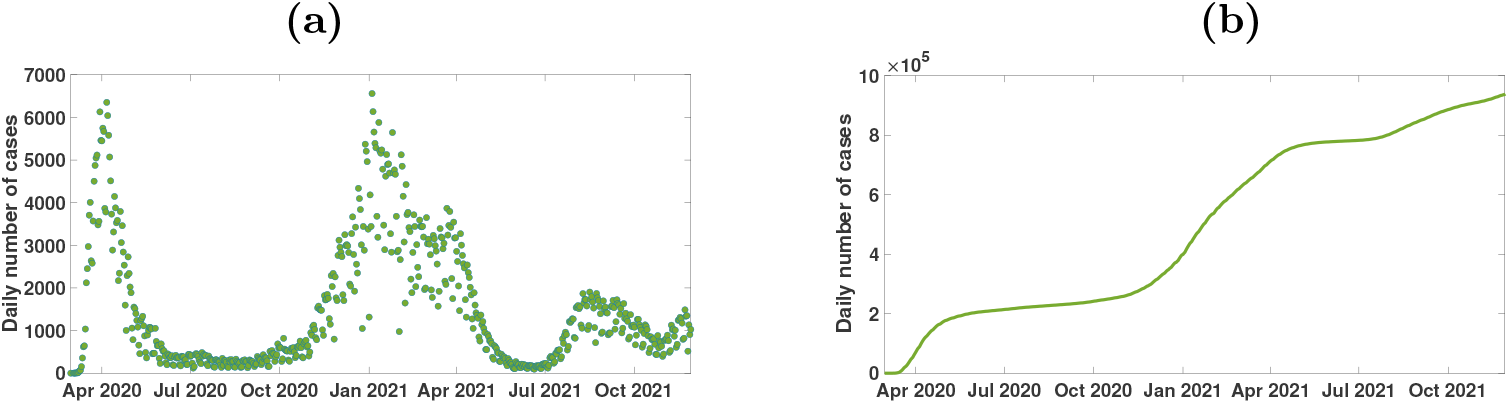
*In (a), we plot* CR_Data_(*t*)′ *the daily number of reported cases of Sars-CoV2 for New York City. In (b), we plot* CR_Data_(*t*) *the cumulative number of reported cases of Sars-CoV2 for New York City*.

In Figure 2-(a), the green dots correspond to the day by day constant values of the function *V*_Data_(*t*) that is used in the model. In Figure 2-(b), the red curve corresponds the function 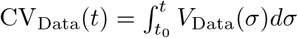 is used in the model.

**Figure 2:**
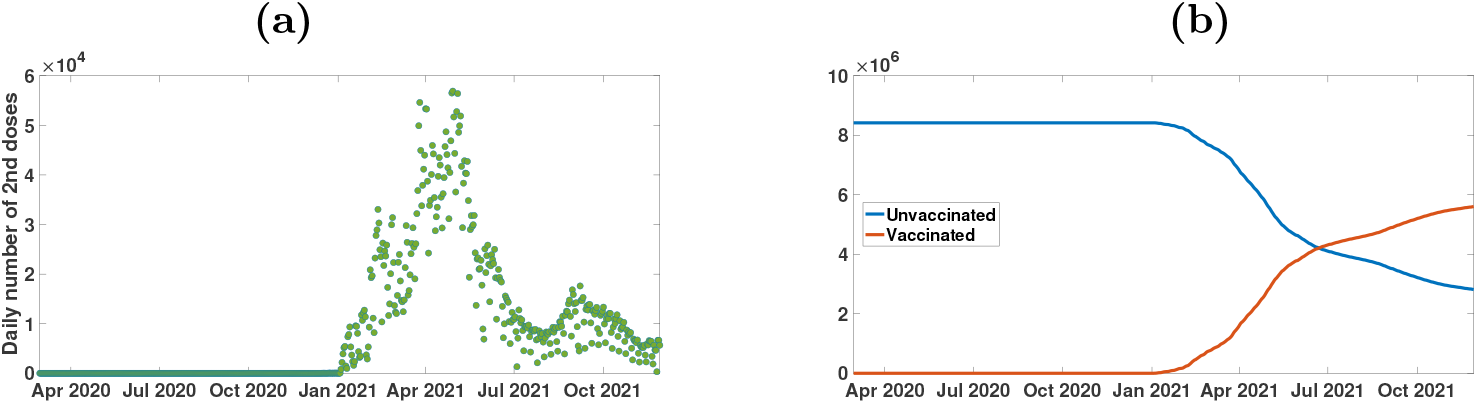
*In (a), we plot V*_Data_(*t*) *the daily number of second doses of vaccine for New York City. In (b), we plot* CV_Data_(*t*) *the cumulative number of second vaccine doses for New York City (red curve), and N* − CV_Data_(*t*) *the number of unvaccinated individuals for New York City (blue curve). The two curves in (b) cross when the number of vaccinated people reaches* 50%.

### 2.2 Epidemic model

Many epidemiological models are based on SIR or SEIR models, which are classical in epidemic modelling. We refer to [24, 20] for early articles devoted to such models and to [1, 2, 5, 3, 4, 6, 8, 17, 21] for later models. In this section, we compare the following SEIUR model to cumulative reported cases data

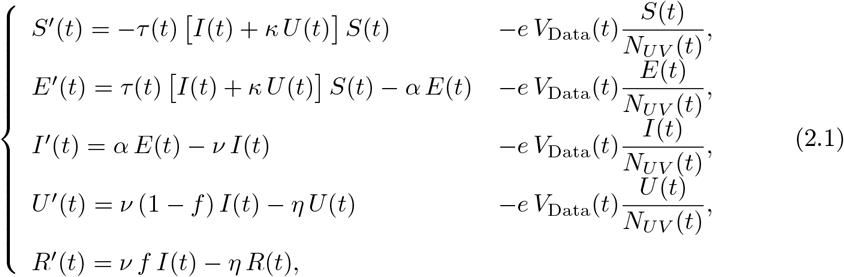

where at time *t, S*(*t*) is the number of susceptible uninfected individuals, *E*(*t*) is the number of exposed individuals (infected, but not yet capable of transmitting the infection), *I*(*t*) is the number of asymptomatic infectious individuals, *R*(*t*) is the number of reported symptomatic infectious individuals, and *U* (*t*) is the number of unreported symptomatic infectious individuals. *N*_*UV*_ (*t*) is the number of unvaccinated individuals. In the model, *S*(*t*)*/N*_*UV*_ (*t*) (respectively, *E*(*t*)*/N*_*UV*_ (*t*), *I*(*t*)*/N*_*UV*_ (*t*), and *U* (*t*)*/N*_*UV*_ (*t*)) is the fraction of susceptible (respectively, infected, reported, and unreported) in the population of unvaccinated individuals.

The system (2.1) is supplemented by the initial data

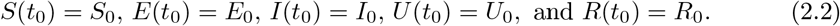

The mathematical model corresponds to the flowchart in Figure 3.

**Figure 3:**
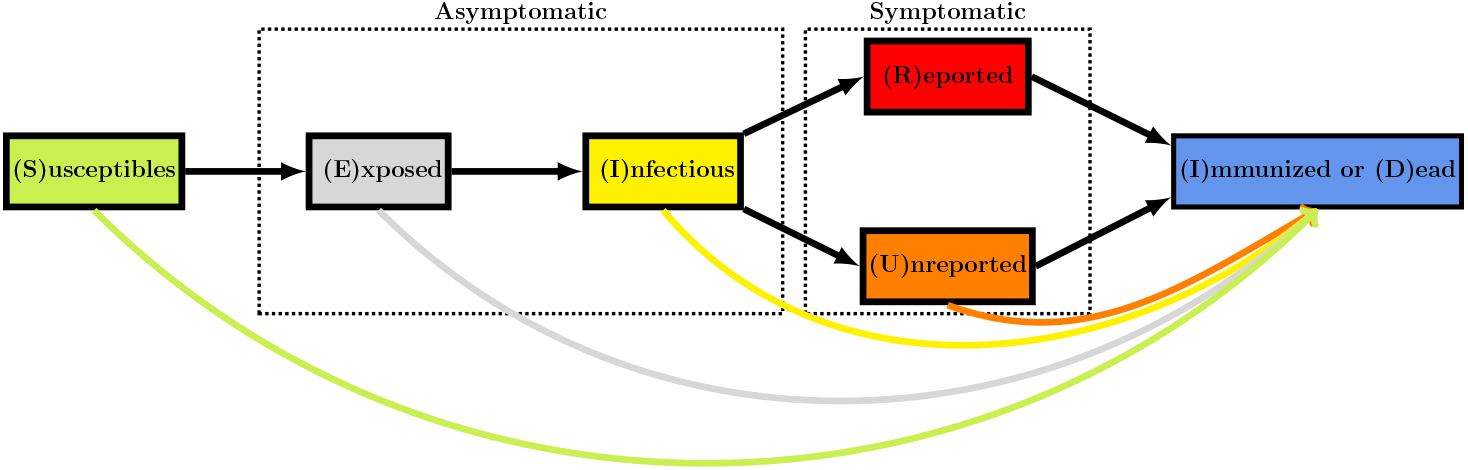
Flowchart for the model. The colored arrows at the bottom represent the vaccination.

In the model, *τ* (*t*) is the time-dependent rate of transmission, 1*/α* is the average duration of the exposed period, 1*/ν* is the average duration of the asymptomatic infectious period, and for simplicity, we subdivide the class of symptomatic infectious individuals into the fraction 0 ≤ *f* ≤ 1 showing severe symptoms, and the fraction 1 − *f* showing mild symptoms, assumed to be undetected. The quantity 1*/η* is the average duration of the symptomatic infectious period for both unreported and reported symptomatic individuals. Asymptomatic infectious and unreported symptomatic infection individuals both contribute to the infection of susceptible individuals, with the parameter *κ* (≥1 or≤ 1) corresponding to their relative contributions. It is assumed that reported symptomatic individuals do not contribute significantly to the transmission of the virus.

In the model, the parameter 0 ≤ *e* ≤ 1 is the vaccine efficacy. This means that when *e* = 0 the vaccine is not effective at all, and if *e* = 1 the vaccine is fully effective. The cumulative number of removed individuals *t* → ID(*t*), immunized (recovered or vaccinated), and or dead, satisfies the equation

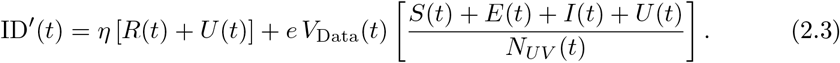

In this model, *V*_Data_(*t*) is the flux of new vaccinated individuals. This means that

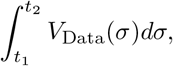

is the total number of vaccinated individuals between *t*_1_ and *t*_2_.

Since no individuals were vaccinated at the start of the epidemic (i.e. for *t* = *t*_0_), we can assume that the total number of individuals *N* in the population at time *t*_0_ is

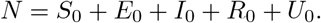

The cumulative number of vaccinated individuals is given by

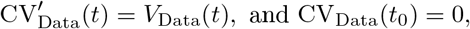

which is equivalent to

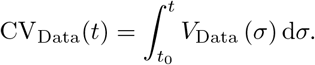

The number of unvaccinated individuals is

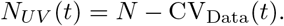

Therefore the model (2.1) can be rewritten as follows

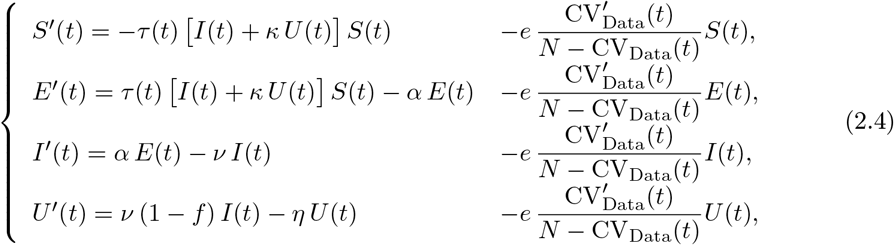

#### Remark 1

*We did not include an R equation in* (2.4), *because the R compartment is decoupled from the rest of the system and we will not use it in the following*.

At the end of the asymptomatic infectious period (corresponding to the *I* compartment), it is assumed that a fraction *f* ∈ (0, 1] of infectious individuals is reported. Therefore, the cumulative number of reported cases CR(*t*) is connected to the epidemic model by the following relationship

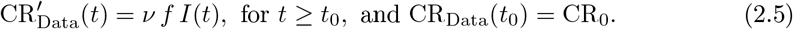

##### Given and estimated parameters

In the model, the data are represented by 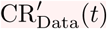, the daily number of reported cases, and *V*_Data_(*t*), the daily number of vaccinations.

In order to compare the model and the data, it is assumed that the known parameters are

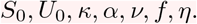

The three remaining parameters are estimated from the above quantities:

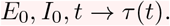

### 2.3 Identification problem

We define the fraction of not effectively vaccinated individuals at time *t*, starting from the time *t*_0_, by

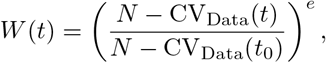

where CV_Data_(*t*) is the cumulative number of second dose vaccinated. Then *W* (0) = 1, and *t* → *W* (*t*) is a non-increasing function. Define

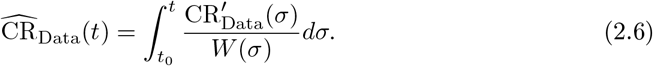

By applying the results in Griette, Demongeot and Magal [11] to the system (A.2) in Appendix A, we obtain the following formula for the rate of transmission expressed in function of the cumulative number of reported cases *t* → CR_Data_(*t*), and the cumulative number of vaccinated individuals *t* → CV_Data_(*t*).

##### Computation of the rate of transmission

The transmission rate is fully determined by the parameters *κ, α, ν, η, f, S*_0_, *E*_0_, *I*_0_, *U*_0_, and by using the five following equations for *t* ≥ *t*_0_

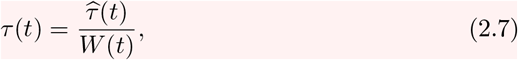

where for *t* ≥ *t*_0_,

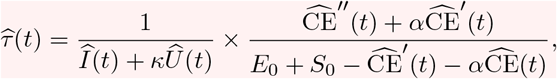

where

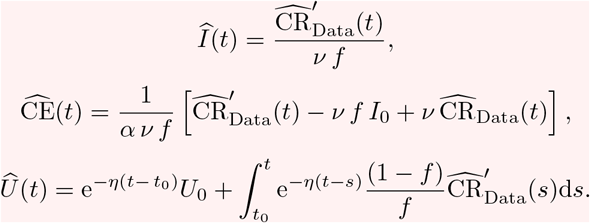

The data that are represented by the functions *t* → CR_Data_(*t*) cumulative number of reported cases, and *t* → *V*_Data_(*t*) the cumulative number of second doses of vaccine are involved in the formula (2.6) to define 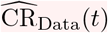.

##### Computation of some initial values from the data

From (2.5) we obtain

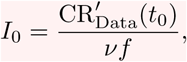

and by using the *I*-equation of system (A.2) and (A.5), we obtain

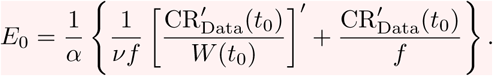

### 2.4 Data normalized by *W* (*t*)

In Figure 4-(a), we plot the daily number of reported cases normalized by *W* (*t*) (the fraction of not efficiently vaccinated individuals at time *t*)

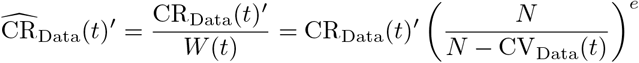

for several values of *e* = 0, 0.25, 0.5,, 0.75, 1.

**Figure 4:**
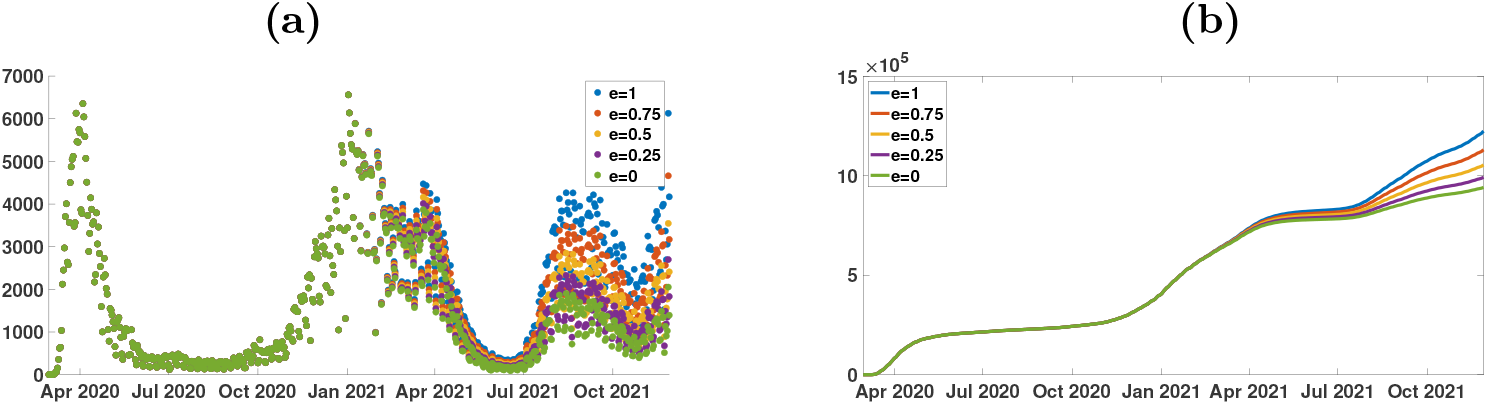
*We plot* 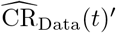 *in (a), and we plot* 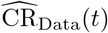 *in (b) for New York City, and e* = 0, 0.25, 0.5,, 0.75, 1. *In (a), the green dots for e* = 0 *corresponds to the original daily number of reported cases* CR_Data_(*t*)′. *In (b), the green curve for e* = 0 *also corresponds to the original cumulative number of reported cases* CR_Data_(*t*).

**Figure 5:**
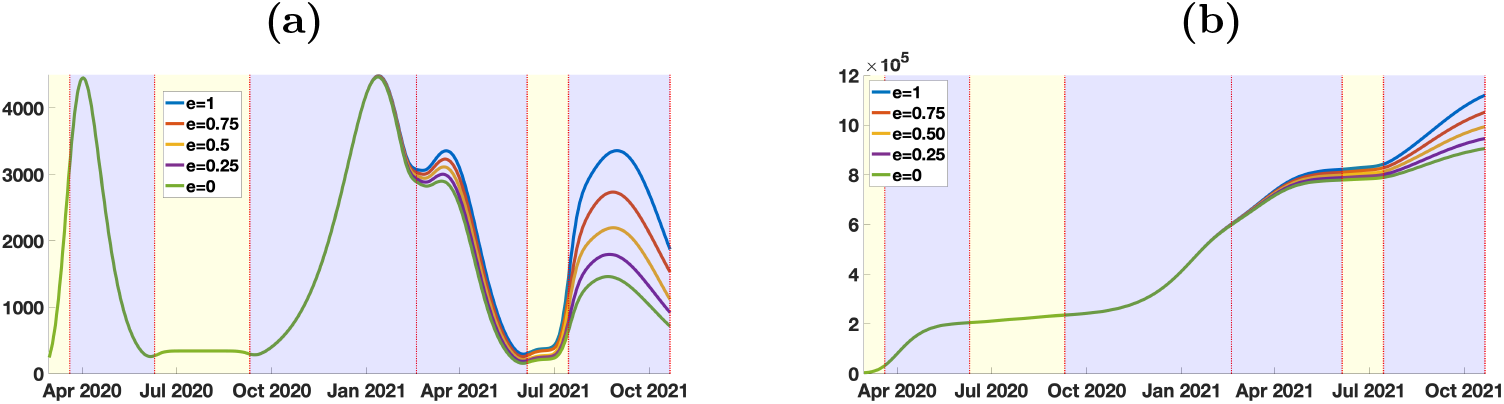
In (a), we plot the phenomenological model used to represent the daily number of reported cases normalized by the efficient vaccinations. In (b), we plot the phenomenological model used to represent the cumulative number of reported cases normalized by the effective vaccinations. The blue background color regions correspond to epidemic phases, and the yellow background color regions to endemic phases.

In Figure 4-(b), we plot the daily number of cases normalized by *W* (*t*), that is

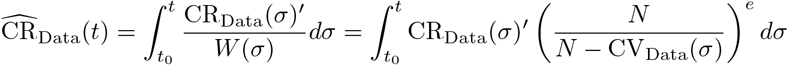

for several values of *e* = 0, 0.25, 0.5,, 0.75, 1.

### 2.5 Phenomenological model

This section is devoted to the phenomenological model used to regularize the data. We refer to [10, 11] for more information. The phenomenological model is fitted to the cumulative reported cases data during the epidemic periods and extended by a lines in between. We regularize the junction point between the period where the phenomenological model has changed. The regularization is obtained using a convolution with a Gaussian function having a standard deviation equal to 7 days.

### 2.6 Instantaneous reproduction numbers

In order to compute the day by day transmission rate *t* → *τ* (*t*), we use the formula (2.7). Then by using the model (A.2) we can consider the problem of the instantaneous reproduction numbers (see [11] for more information). To investigate the role of the vaccination for the COVID-19 data, we use our method to compute the transmission rate, and we consider the **instantaneous reproduction number with vaccination**

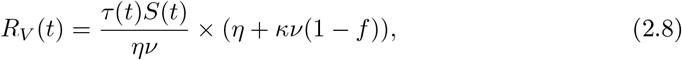

the **quasi-instantaneous reproduction number with vaccination**

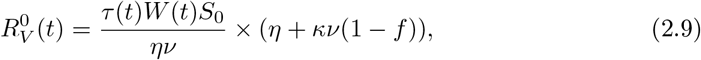

and the **quasi-instantaneous reproduction number without vaccination**

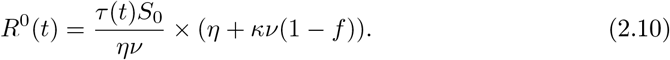

## 3 Results

The parameters used in the simulations are listed in Table 1 in Appendix B.

**Table 1:**
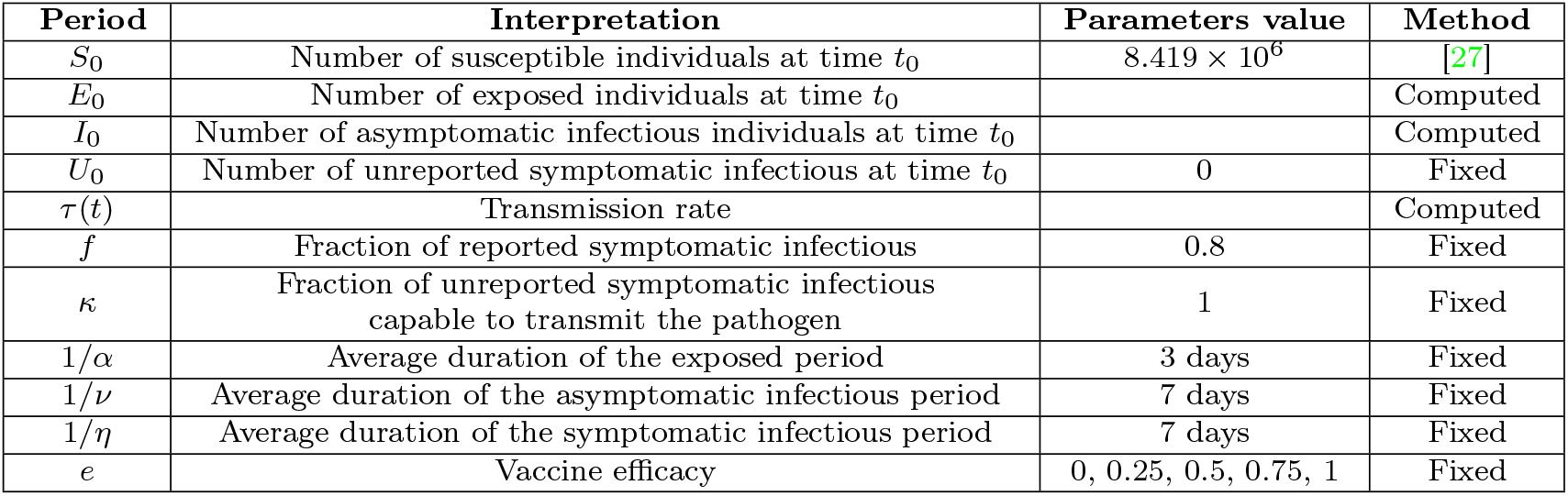
In this table we list the values of the parameters of the epidemic model used for the simulations.

In Figure 6, we observe almost no influence of the vaccine efficacy *e* on the basic reproduction number. This is due to some compensatory effects between *τ*(*t*) and *S*(*t*), because *τ*(*t*) and *S*(*t*) are evaluated to adjust the number of cumulative reported cases, which is fixed.

**Figure 6:**
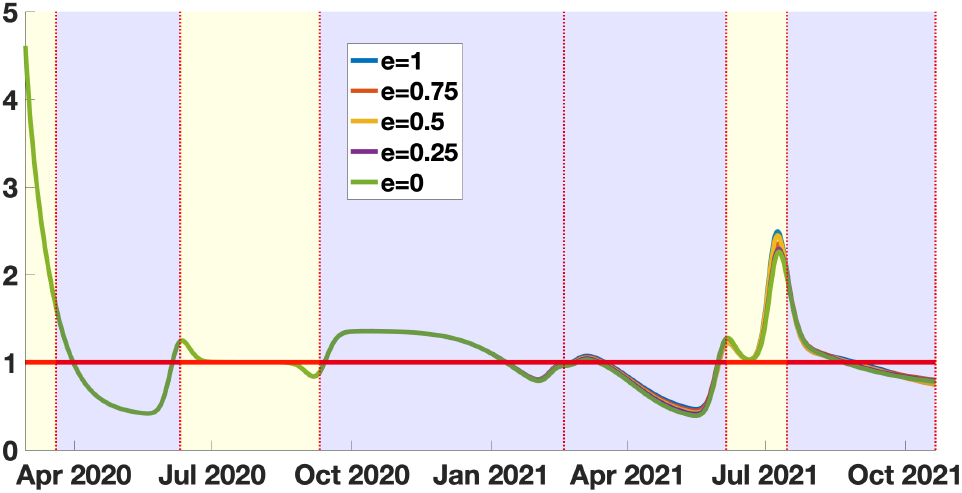
In this figure we plot the instantaneous reproduction number with vaccination 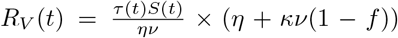. The blue background color regions correspond to epidemic phases, and the yellow background color regions to endemic phases.

In Figure 7, we see almost no difference with Figure 6. It means that the cumulative number of infected is so small compared to the total size of the population of New York City that the 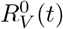 is almost unchanged compared to *R*_*V*_(*t*). This means that the cumulative number of infected is too small to have a significant impact to reduce the basic reproduction number.

**Figure 7:**
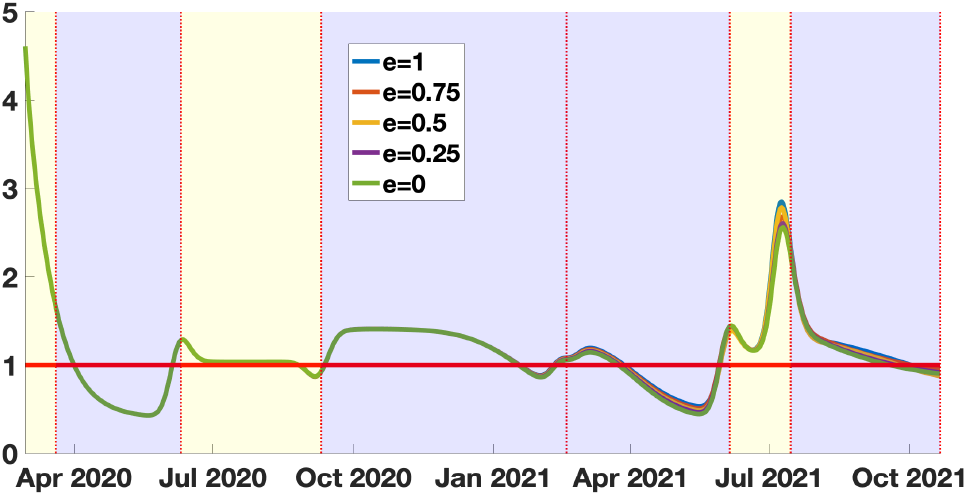
In this figure we plot the quasi-instantaneous reproduction number with vaccination 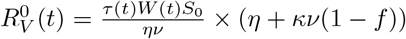. The blue background color regions correspond to epidemic phases, and the yellow background color regions to endemic phases.

In Figure 8, the blue curve corresponds the *R*^0^(*t*). The instantaneous reproduction number should be interpreted as the instantaneous reproduction number in the absence of vaccination, conditionality to the fact that the vaccine is fully efficient. The same interpretation holds for *e* = 0.75, 0.5, 0.25, 0. This means that vaccination has a strong influence on the dynamic of the epidemic. This influence indeed strongly depends on the vaccine efficacy *e*. We can see that during the most recent epidemic wave, the situation in New York City would have been much worst in the absence of vaccination. We have indeed, for *e* = 0.75, the last peak of the red curve representing *R*^0^(*t*) around 4.5.

**Figure 8:**
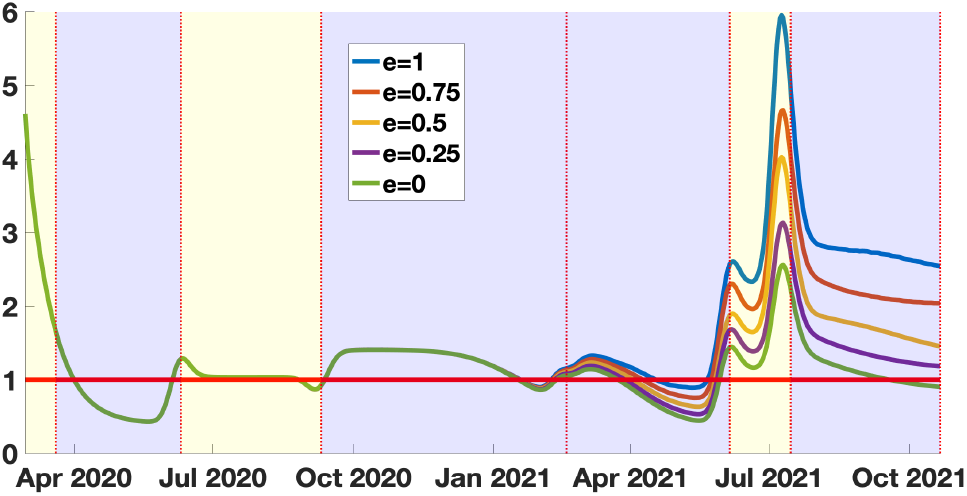
In this figure we plot the quasi-instantaneous reproduction number without vaccination 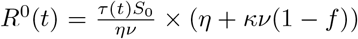. The blue background color regions correspond to epidemic phases, and the yellow background color regions to endemic phases. One may observe that R^0^(t) is a multiple of the time dependent transmission *rate*.

We cannot estimate the value of the vaccine efficacy, because for each value of *e*, we obtain a perfect match with the data (i.e. a perfect correspondence with the phenomenological model plotted in Figure 5-(b)). But conditionally to the value of *e*, we can draw some conclusions. If the vaccine efficacy is above 0.75, it means that, during the last epidemic wave, New York City escaped an epidemic wave as bad (or worse) than the first one (this corresponds to the region between the blue curve and the red curve in Figure 8 in the last blue background color regions). If the vaccine efficacy is between 0.25 and 0.75, we can see a significant gain compared to the green curve (this corresponds to the region between the purple curve and red curve in Figure 8 in the last blue background color regions).

The above result means that a loss of vaccine efficacy increases the number of susceptible patients. So that with an equal number of daily reported cases, the reproduction number must decay. Conversely, if the vaccine efficacy increases, the reproduction number must increase.

## 4 Discussion

In this article, we developed a new method to model the COVID-19 epidemic by using the daily reported cases and vaccination data. We use phenomenological models to get an exploitable reconstruction of the history of the epidemic and develop a new method to identify the parameters of an epidemic model with vaccination that reproduces the exact behavior of the data.

Since the first efficient vaccines against SARS-CoV-2 appeared at the end of 2020, many countries have implemented vaccination policies to protect their population. As a result, a non-negligible fraction of the population has acquired at least a partial immunity against the disease. This means that the number of susceptible hosts has been significantly reduced. Several studies, including some authors’ works, developed methods to connect the data with epidemic models. In the presence of vaccination, these models may over-estimate the number of susceptible hosts, and their conclusions should therefore be taken with precaution. We correct this flaw by including vaccination data in our model in the present study. We construct, in particular, the transmission rate *τ* (*t*) and the instantaneous reproduction number *R*_*V*_ (*t*) of the disease.

In Figures 6 and 7, we present our computations concerning the instantaneous and quasi-instantaneous reproduction number *R*_*V*_ (*t*) and 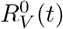. The explicit formula developed in this paper allows us to investigate the role of the vaccine efficacy parameter *e*. Surprisingly, the instantaneous reproduction number reconstructed from the data does not depend very much on the vaccine’s efficacy. We observe that the five curves presented in Figure 6 are almost equal (the same can be said about the quasi-instantaneous reproduction number in Figure 7). We understand this phenomenon as a balance between the number of susceptible hosts *S*(*t*) and the transmission rate *τ* (*t*). This is because, since the data is unchanged, the increase in the efficacy of the vaccine reduces the number of available hosts, so the transmission rate must be increased to recover the data. In this process, the product *τ* (*t*)*S*(*t*) (and therefore *R*_*V*_ (*t*) and 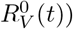 is almost unchanged.

Immunization data can help to understand better the effectiveness of a city, a state, or a country’s immunization policy. Strictly speaking, this policy should be adapted to the populations at risk and, for example, be different according to the age groups and take into account the progressive degradation of the immunity conferred by the vaccines, due to the appearance of variants of the initial virus.The data used in this article do not consider any specificity of sub-populations for New York City. Therefore the age groups were not considered, nor the extinction of immunity over time, which could be taken into account by considering an efficacy *e* dependent on the time and a flux of the vaccinated back into the susceptible compartment.

Despite these shortcomings, the model clearly shows the impact of the vaccine policy on the epidemic dynamics, thanks to the explicit formulas allowing the calculation of important parameters, such as the transmission rate. In addition, the model allows for the introduction of additional elements, when documented by observed data, such as age, loss of immunization, and cross-immunization. This last phenomenon causes weights on the vaccine policy’s effectiveness, which is interesting for further investigation.

Vaccination confers a new immunity, which is in addition to a possible pre-existing cross-immunity [19], and the vaccination policy could be adjusted in relation to the response via cross-immunity against epitopes common to numerous coronaviruses. For example, if the age classes are considered, young individuals are those whose cross-immunity is still active, causing a strong response to vaccination with possible systemic undesirable effects. Consequently, it would be interesting in the future to develop improvements of the model in order to refine the number of doses per target population at risk and thus ensure, for a smaller quantity of vaccinated, the same efficacy in the immunization of the general population.

Our model could be extended in several other directions. Here we did not distinguish between immunized, dead, and vaccinated individuals. These could be added to the model, and other phenomena could be included as well, like a different fading rate of immunity coming from the disease and the vaccine, provided the associated parameters (death rate, etc.) are known. By including age classes, we could also distinguish the strength of the immune response according to age and better measure the benefit-risk ratio of vaccination with respect to age. We could also implement a different rate of loss of immunity by age class.

## Data Availability

No data were produced in this study

## Appendix

### A Transformation of the system into a system into a standard epidemic model

The goal of this section is to connect the model with vaccination to the model without vaccination used previously in [11]. Our goal is to apply to the transformed system some of the results obtained in [11]. Set

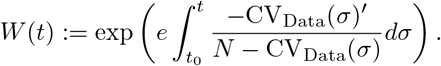

Then

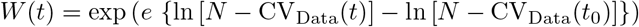

therefore

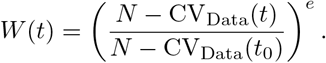

By integrating the *S*-equation of system (2.4) we obtain

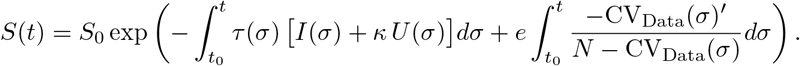

Hence we have

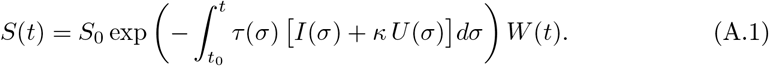

Define

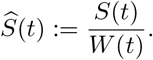

It follows that from (A.1) that

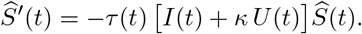

By setting

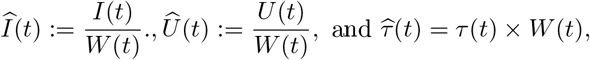

we obtain

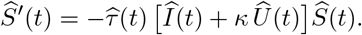

By replacing (A.1) in the *E*-equation of system (2.4), we obtain

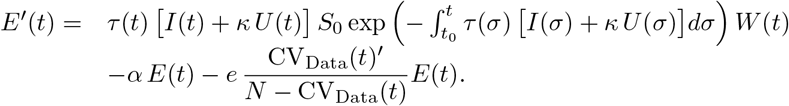

We observe that

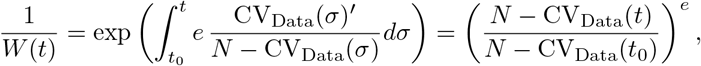

and

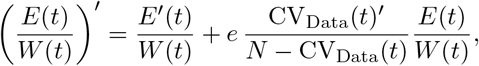

therefore, we obtain

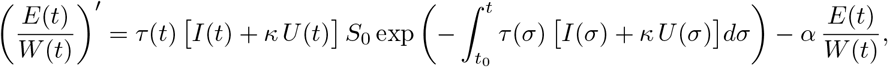

and by setting

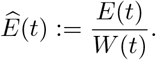

we obtain

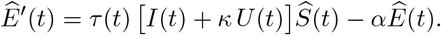

By using similar argument for the remaining equations of (2.4) we obtain the following result.

#### Lemma 2

**(Transformation of the system)** *We define for t* ≥ *t*_0_,

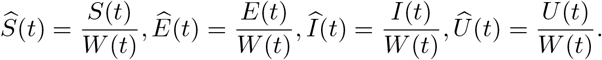

*Then the system* (2.4) *becomes for each t* ≥ *t*_0_,

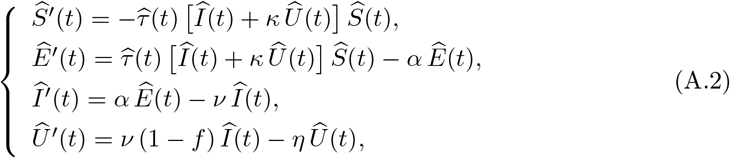

*with initial data*

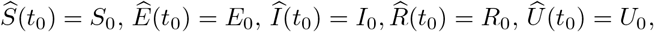

*and*

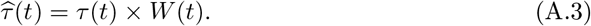

#### Connection with the data

By using the equation (2.5) (connecting the model and the data), we obtain

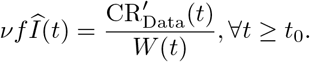

We define for all *t* ≥ *t*_0_,

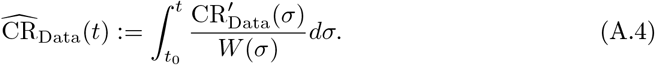

Then we obtain the same formula than (2.5) (with hat), namely

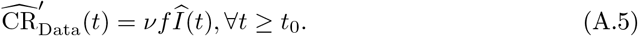

### B Table of parameters

## References

[1] Anderson, R. M., and May, R. M., Infectious Diseases of Humans: Dynamics and Control, Oxford University Press, 1992.

[2] Bailey, N. T. J., The Mathematical Theory of Epidemics, Hafner Publishing Co., New York, 1957.

[3] Brauer, F., and Castillo-Chavez, C., Mathematical Models in Population Biology and Epidemiology, Springer, New York, 2nd edition, 2012.

[4] Brauer, F., Castillo-Chavez, C. and Feng, Z., Mathematical Models in Epidemiology, Springer, New York, 2019.

[5] Brauer, F., van den Driessche, P., and Wu, J., Mathematical Epidemiology, Springer, Berlin, Germany, 2008.

[6] Busenberg, S., and Cooke, K., Vertically Transmitted Diseases, Springer-Verlag, Berlin, 1993.

[7] Demongeot, J., Griette, Q., and Magal, P., SI epidemic model applied to COVID-19 data in mainland China. Royal Society Open Science 7:201878 (2020).

[8] Diekmann, O., Heesterbeek, H., and Britton, T., Mathematical Tools for Understanding Infectious Disease Dynamics, Princeton University Press, Princeton, NJ, 2013.

[9] Diekmann, O., Heesterbeek, J. A. P., and Metz, J. A. J., On the definition and the computation of the basic reproduction ratio r<sub>0</sub> in models for infectious diseases in heterogeneous populations, J. Math. Biol., 28 (1990), 365–382. doi: 10.1007/BF00178324.

[10] Griette, Q., Demongeot, J., and Magal, P., A robust phenomenological approach to investigate COVID-19 data for France. Mathematics in Applied Sciences and Engineering, 2(3) (2021), 149–218.

[11] Griette, Q., Demongeot, J., and Magal, P., What can we learn from COVID-19 data by using epidemic models with unidentified infectious cases? Mathematical Biosciences and Engineering, 19(1) (2021), 537?594.

[12] Griette, Q., and Magal, P., Clarifying predictions for COVID-19 from testing data: The example of New York State, Infectious Disease Modelling, 6 (2021), 273–283.

[13] Iboi, E. A., Ngonghala, C. N., and Gumel, A. B., Will an imperfect vaccine curtail the COVID-19 pandemic in the US? Infectious Disease Modelling, 5 (2020), 510–524.

[14] Li, Q., Tang, B., Bragazzi, N. L., Xiao, Y., and Wu, J., Modeling the impact of mass influenza vaccination and public health interventions on COVID-19 epidemics with limited detection capability. Mathematical biosciences, 325, 108378 (2020).

[15] Moore, S., Hill, E., Dyson, L., Tildesley, M., and Keeling, M., Modelling optimal vaccination strategy for SARS-CoV-2 in the UK, PLOS Computational Biology, 17(5) (2021).

[16] Moore, S., Hill, E., Tildesley, M., Dyson, L., and Keeling, M., Vaccination and non-pharmaceutical interventions for COVID-19: a mathematical modelling study, The Lancet Infectious Diseases, 21(6) (2021), 793–802.

[17] Murray, J. D., Mathematical Biology, Springer-Verlag, Berlin, 1989.

[18] Perra N., Non-pharmaceutical interventions during the COVID-19 pandemic: A review. Physics Reports (2021).

[19] Simula, E.R., et al., HCoV-NL63 and SARS-CoV-2 share recognized epitopes by the humoral response in sera of people collected pre-and during CoV-2 pandemic. Microorganisms, 8(12) (2020), 1993.

[20] Tang, B., et al., Estimation of the transmission risk of the 2019-nCoV and its implication for public health interventions, J. Clin. Med., 9 (2020), 462. doi: 10.3390/jcm9020462.

[21] Thieme, H.R., Mathematics in Population Biology, Princeton University Press, Princeton, NJ, 2003.

[22] Webb, G., A COVID-19 epidemic model predicting the effectiveness of vaccination, Mathematics in Applied Sciences and Engineering, (2021), 1–15.

[23] Webb, G., A COVID-19 epidemic model predicting the effectiveness of vaccination in the US, Infectious Disease Reports, 13(3) (2021), 654–667.

[24] Wu, J. T., et al., Estimating clinical severity of COVID-19 from the transmission dynamics in Wuhan, China, Nat. Med., 26 (2020), 506–510. doi: 10.1038/s41591-020-0822-7.

[25] Yahi, N., Chahinian, H., and Fantini J., Infection-enhancing anti-SARS-CoV-2 antibodies recognize both the original Wuhan/D614G strain and Delta variants. A potential risk for mass vaccination?. Journal of Infection (2021).

[26] New York City Department of Health and Mental Hygiene https://www1.nyc.gov/site/doh/covid/covid-19-data.page (accessed on 17 December 2021).

[27] United States census bureau https://www.census.gov/en.html (accessed on 17 December 2021).

